# Hypertension care cascade in Nepal: findings from Nepal Demographic and Health Survey 2022

**DOI:** 10.1101/2025.03.10.25323662

**Authors:** Ashok Khanal, Sulochan GC, Irusha Dahal, Sumi Mishra, Vijay S. GC, Sharada Prasad Wasti, Rakesh Ghimire

## Abstract

**Background:** Hypertension is a leading risk factor to Nepal’s rising burden of cardiovascular diseases, yet many affected individuals remain undiagnosed, untreated, and uncontrolled. Identifying gaps in care and variations across socio-demographics and provinces can help optimize interventions to prevent and control hypertension.

**Objectives:** We aimed to quantify the prevalence and gaps in hypertension awareness, treatment, and control, as well as their determinants, using the latest nationally representative data from the 2022 Nepal Demographic and Health Survey (NDHS).

**Methods:** We used the NDHS 2022, conducted from January 5 to June 22, 2022. Socio-demographic factors such as sex, education level, age, body mass index (BMI), marital status, and residency were used to examine hypertension care cascade metrics. Logistic regression was used to assess the factors associated with each outcome above.

**Results:** A total of 9,990 unweighted observations, representing 10,065 participants (4,321 males and 5,744 females) aged ≥15 years were included in this study. The national prevalence of hypertension was 20⸱0%. In multivariate analysis, male sex, older age, lower education level, married/divorced individuals, higher BMI, and urban residence had increased odds of hypertension. Among hypertensive patients, 50⸱2% were aware of their diagnosis, 31⸱7% were receiving treatment, and 18⸱0% had controlled blood pressure. There were substantial variations across the seven provinces in hypertension prevalence, ranging from (14⸱2%) in Karnali to (25⸱5%) in Koshi. The unmet need for hypertension diagnosis, treatment, and control was highest in Sudurpaschim province.

**Conclusion:** In this cross-sectional survey study, about one out of every five Nepalese are hypertensive. Furthermore, the gaps in the hypertension care cascade are huge, with over 80% of adults with hypertension are either undiagnosed, untreated, or treated but with uncontrolled hypertension. Targeted and de-centralized improvements in access to early hypertension diagnosis and affordable treatment are especially crucial for low-income households, remote areas, and younger populations.

## Introduction

Like many low- and middle-income countries (LMICs), Nepal is experiencing a significant shift in its disease burden, marked by a substantial rise in non-communicable diseases (NCDs)(1–4). This concerning transition can be attributed to a confluence of factors, including economic development, rapid urbanization, changing dietary patterns and physical activity levels (5), and an ageing population (driven by increased life expectancy) (6). The Global Burden of Disease Study 2019 underscores this alarming trend, with the proportion of deaths attributed to NCDs skyrocketing from 31.3% in 1990 to a concerning 71.1% in 2019 (7). Notably, cardiovascular disease (CVD) was the leading cause of death, with 24% of total deaths being attributable to CVDs (7). Furthermore, hypertension, a well-established CVD risk factor, has been identified as the second leading risk factor of total deaths in Nepal (4,7), surpassing traditional threats like malnutrition, vaccine-preventable diseases, and tobacco use.

Antihypertensive medications are inexpensive and effective, yet only a minority of adults with hypertension in Nepal are diagnosed and treated (8,9). To address this critical gap and rising cardiovascular disease (CVD), Nepal launched the Hypertension Care Cascade Initiative (10). Backed by all government levels, it aims to improve detection and management within the nation’s primary healthcare system, targeting 1.5 million people with hypertension and diabetes for protocol-based treatment by 2025 (10). However, a crucial gap remains—the lack of up-to-date data on hypertension care gaps to identify areas needing intervention and benchmarking progress in blood pressure control both at national and sub-national levels.

While existing research extensively explores the prevalence and determinants of hypertension in Nepal (11–13), a critical gap exists in our understanding of current awareness, treatment, and control rates. Prior studies examining the hypertension care continuum either relied on pre-2020 data (8,14,15), lacked provincial breakdowns (8,14,15), or focused on specific populations (16–18). Up-to-date regional data may, therefore, empower local governments to optimize the planning, implementation, and monitoring of investments for the Hypertension Care Cascade Initiative in Nepal (launched in 2023).

We utilize the “cascade of care” approach to pinpoint where in the hypertension care continuum patients are lost and how these patterns vary between different socio demographics and provinces. This method helps healthcare providers and stakeholders identify gaps in hypertension control, enabling the pairing of at-risk populations with the most appropriate interventions (e.g., prioritizing diagnosis, promoting medication adherence, and improving retention programs). Using data from a nationally representative sample of Nepalese adults (15 to 95 years old), this study, therefore, aims to describe the comprehensive picture of the current state of hypertension care across Nepal - prevalence, diagnosis, treatment, and control - at both national and provincial levels.

## Methods

### Data source

This study analyzed secondary data from the sixth Nepal Demographic and Health Survey (19)(NDHS) 2022, a nationally representative cross-sectional survey, to describe the hypertension care cascade at both national and provincial levels. Conducted as part of the DHS program by New ERA under the guidance of the Ministry of Health, Government of Nepal, and supported by ICF International and the United States Agency for International Development (USAID), the primary objective of the 2022 NDHS was to provide up-to-date estimates of basic demographic and health indicators.

The 2022 NDHS utilized a two-stage stratified sampling method. The seven provinces were divided into urban and rural areas for stratification. In the first stage, 476 primary sampling units (PSUs) were selected based on probability proportional to PSU size, with 248 PSUs from urban areas and 228 from rural areas. In the second stage, 30 households were selected from each PSU, resulting in a total sample size of 14,280 households—7,440 urban and 6,840 rural. Blood pressure measurements were conducted on all women and men aged 15 years or older in a subset of selected households, constituting one-fourth of the total households selected for the 2022 NDHS (and half of the households selected for biomarker information collection). A total of 4,601 men and 6,021 women aged 15 years or older were eligible for blood pressure measurements. Of these, 96% of women and 92% of men had their blood pressure measured and almost all had three measurements. Detailed methodology of the NDHS is available in the most recent NDHS report (19). ***Figure 1 in Supplement*** presents the details of the sample size and exclusion criteria for the population analyzed to describe the hypertension care cascade.

**Figure 1:**
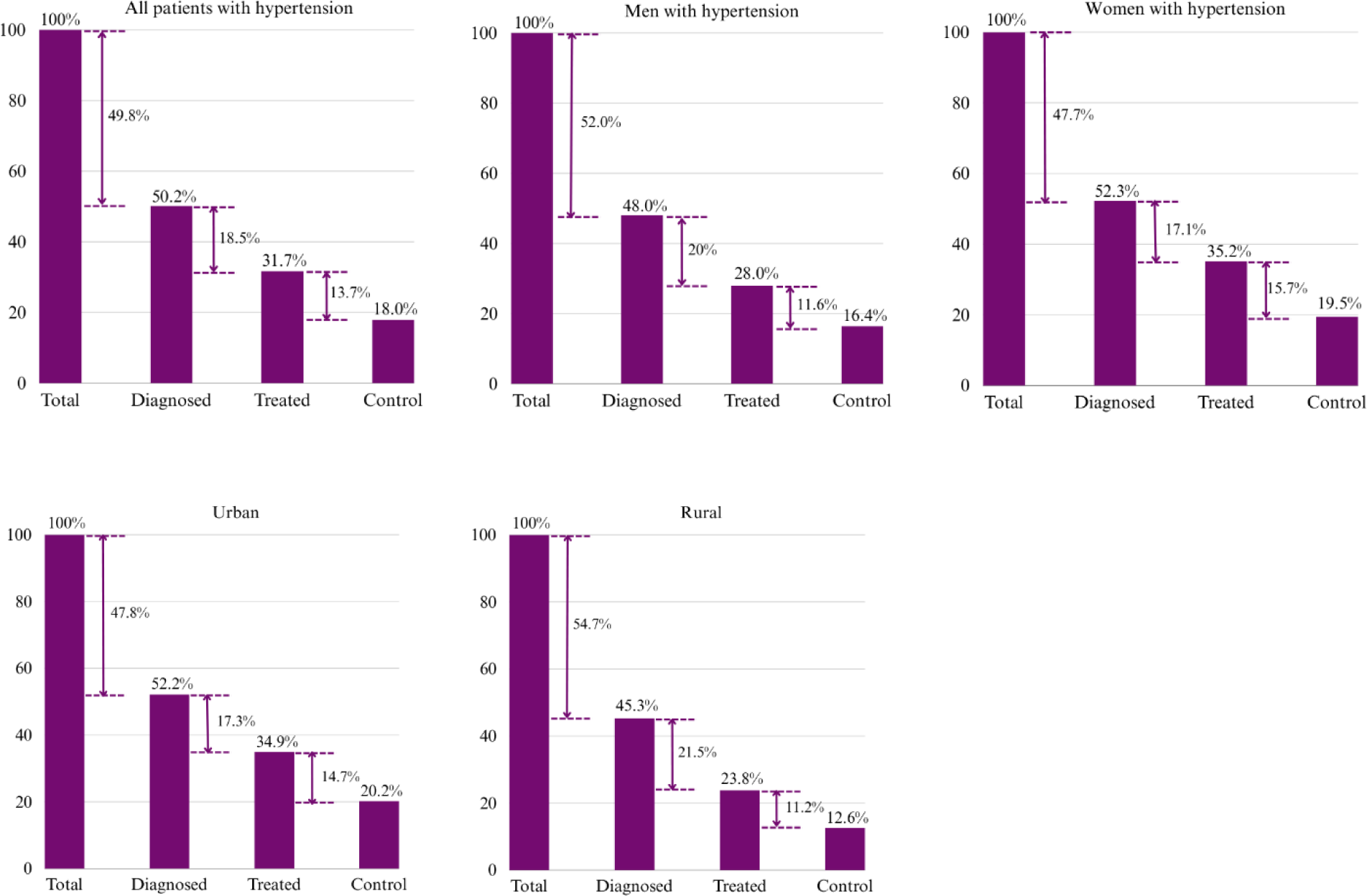
Hypertension care cascade in Nepal. The numbers between bars indicate the percentage loss at each stage of the cascade. Total = total participants diagnosed with hypertension at the time of survey.

### Data collection

The Nepal Demographic and Health Survey (NDHS) 2022 was conducted from January 5 to June 22, 2022, by 19 field teams, each comprising a supervisor, one male interviewer, three female interviewers, and one biomarker specialist (19). The survey employed a standardized tool validated globally and refined for the Nepali context through expert consultations, which had been reliably used in previous periodic surveys. Rigorous procedures ensured prompt addressing data quality issues like incompleteness, inconsistencies, and errors. Regarding blood pressure measurement, trained examiners followed standardized protocols using a validated automatic upper arm monitor (A&D Medical, Tokyo) with three cuff sizes (medium, small, extra-large) chosen based on the participant’s bare upper arm circumference. Three readings were taken at ≥ 5-minute intervals, with the mean of the last two determining the final blood pressure level. Nearly all participants had three measurements recorded. If the third reading was unavailable, the second reading alone categorized hypertension status; similarly, if both the second and third were missing, the first reading alone determined the classification.

### Hypertension Care Cascade: Diagnosis, Treatment, and Control

Hypertension was defined as a systolic blood pressure of ≥140 mmHg and/or a diastolic blood pressure of ≥90 mmHg and/or the current use of medication prescribed for hypertension (20,21). Participants were classified as aware or diagnosed if they met the hypertension definition mentioned above and reported having ever been diagnosed with hypertension by a doctor or health worker. Patients were considered treated if they reported currently receiving medication for hypertension. Additionally, patients receiving medication with a systolic blood pressure <140 mmHg and diastolic blood pressure <90 mmHg were classified as having controlled blood pressure.

We calculated the treated proportion separately among the overall sample with hypertension, regardless of diagnosed status for hypertension, and among those diagnosed. Similarly, we calculated the controlled proportion separately for the overall sample with hypertension, regardless of treatment status for hypertension, and among those receiving treatment. The definitions are summarized in Table 1 in Supplement.

**Table 1:**
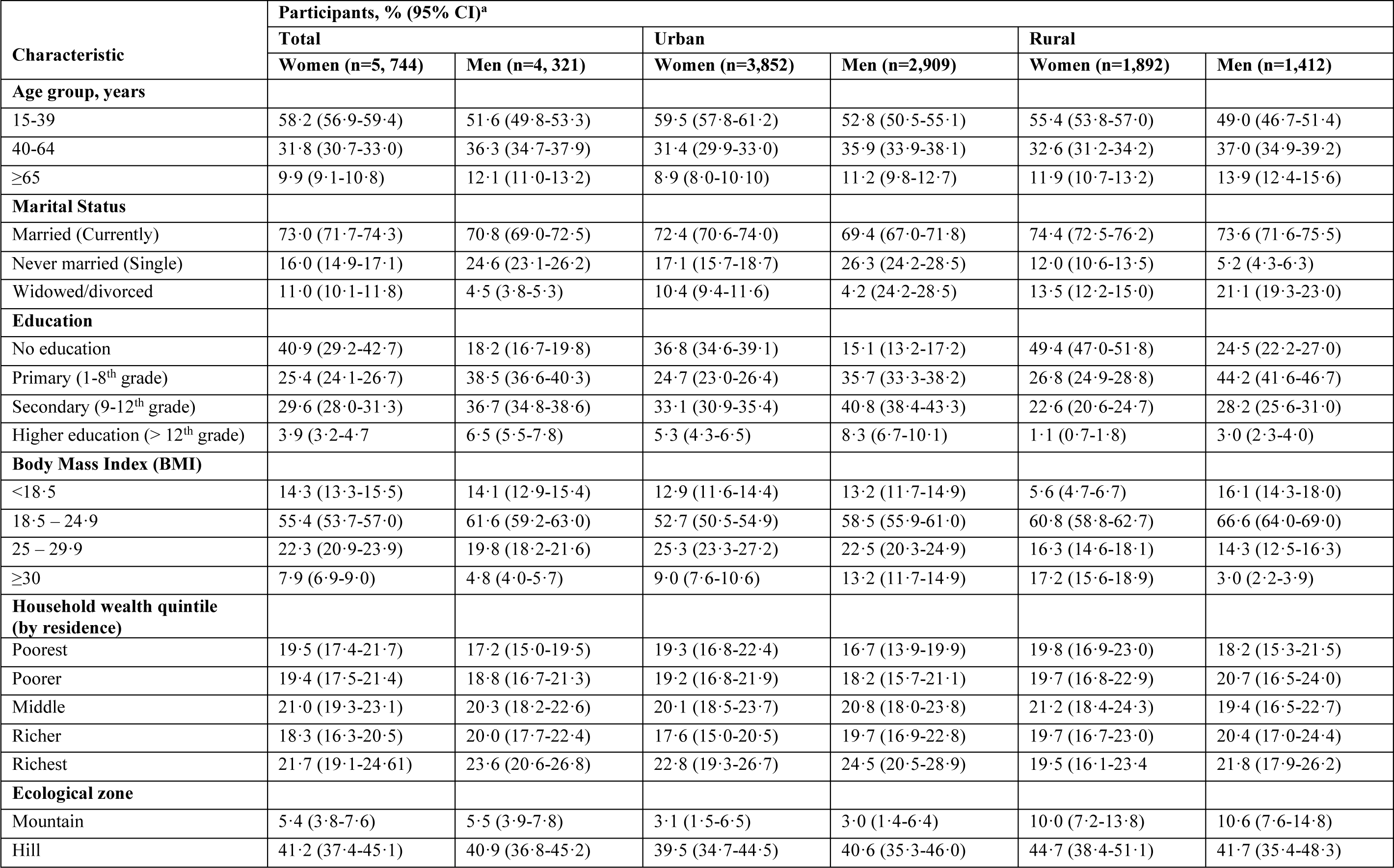
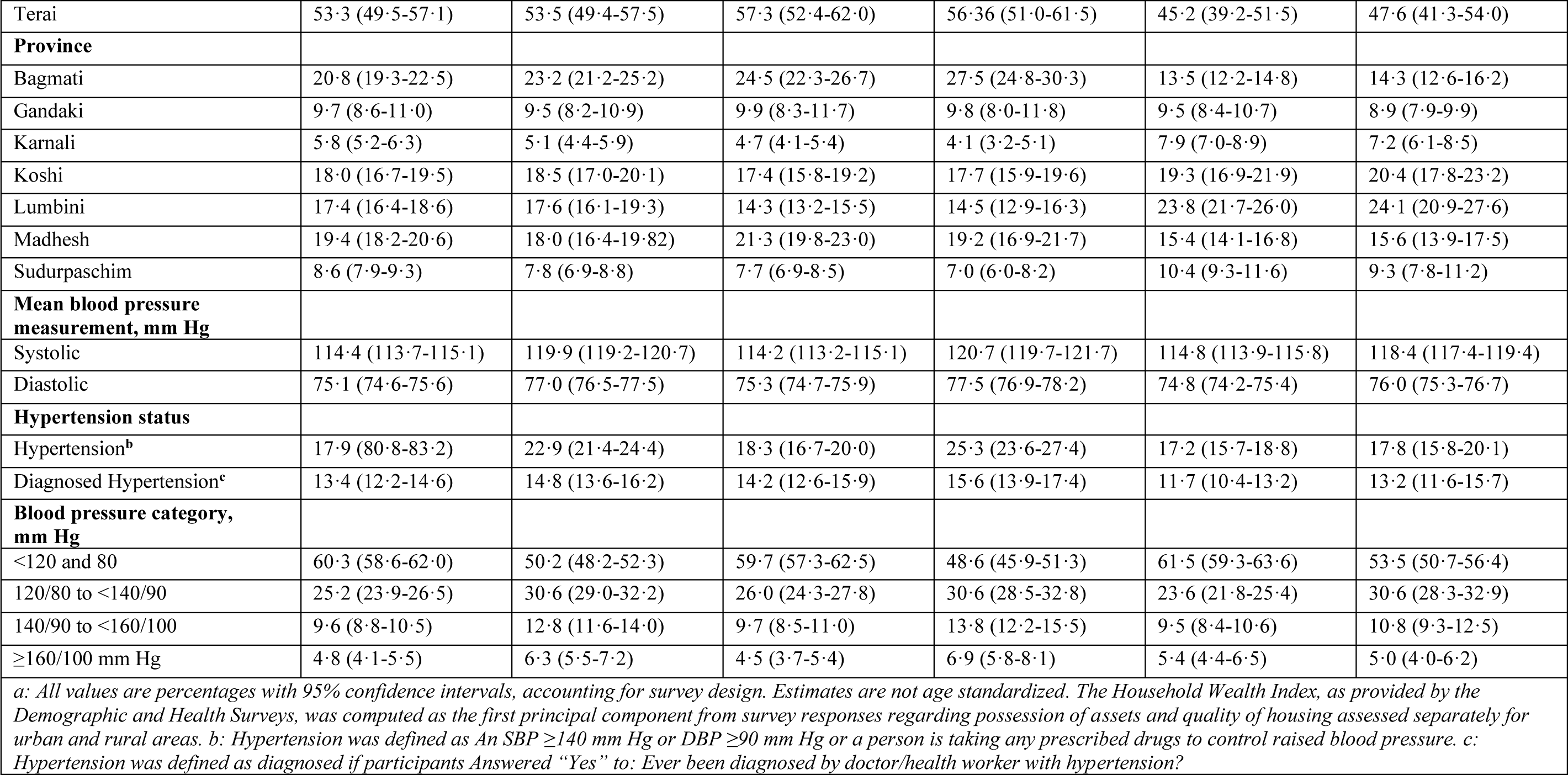
Characteristics of Participants in the Analytic Sample for Estimating Care Cascade of Hypertension in Nepal (N =10, 065)

### Predictors of reaching each care cascade step

We examined hypertension care cascade metrics by five individual-level sociodemographic factors: sex (male or female), and education level (none, basic [up to 8th grade], secondary [up to 12th grade], or higher [above 12th grade]), age (15-39, 40-64, or 65 years), BMI ([<18[underweight], 18–24.9[normal weight], 25–29.9[overweight], and ≥30[obese]) and marital status single married or formerly married. Additionally, we stratified the analysis based on three household sociodemographic factors: residence (rural or urban), ecological region (mountain, hill, or terai), and regional wealth quintile (urban and rural) using the household wealth index from the DHS. The definitions are summarized in ***Table 2 in Supplement*.**

**Table 2:** Survey weighted prevalence, diagnosis, treatment, and control of hypertension by socio-demographic factors in Nepal.

| Variable | Number of participants | Hypertensives | Prevalence of hypertension<br>(%, 95% CI) | Diagnosed hypertension<br>(aware)<br>(%, 95% CI) | Treated | Among Diagnosed<br>(%, 95% CI) | Controlled | Among Treated<br>(%, 95% CI) |
| --- | --- | --- | --- | --- | --- | --- | --- | --- |
|  |  |  |  |  | Among all<br>hypertensives<br>(%, 95% CI) |  | Among all<br>hypertensives<br>(%, 95% CI) |  |
| **Total** | 10065 | 2019 | 20.0 (19.0-21.2) | 50.2 (47.3-53.2) | 31.7 (28.8-34.7) | 63.2 (59.4-66.8) | 18.0 (15.5-20.8) | 56.7 (51.2-62.0) |
| **Sex** |  |  |  |  |  |  |  |  |
| Female | 5744 | 1031 | 17.9 (16.7-19.2) | 52.3 (48.5-56.2) | 35.2 (31.5-39.1) | 67.3 (62.6-71.6) | 19.5 (16.5-22.9) | 55.3 (48.8-61.5) |
| Male | 4321 | 988 | 22.9 (21.4-24.4) | 48.0 (44.1-51.9) | 28.0 (24.6-31.82) | 58.5 (53.1-63.6) | 16.4 (13.3-20.1) | 58.6 (50.4-66.3) |
| **Age group, years** |  |  |  |  |  |  |  |  |
| 15-39 | 5571 | 404 | 7.2 (6.4-8.2) | 31.5 (25.9-37.7) | 10.8 (7.8-14.7) | 34.3 (24.8-45.3) | 7.8 (5.2-11.5) | 71.9 (55.2-84.1) |
| 40-64 | 3400 | 1096 | 32.2 (30.3-34.2) | 53.1 (49.5-56.7) | 33.2 (29.8-36.8) | 62.6 (57.9-67.1) | 18.7 (15.7-22.2) | 56.3 (49.3-63.1) |
| ≥65 | 1094 | 519 | 47.4 (43.9-51.0) | 58.7 (53.1-64.2) | 44.8 (38.7-51.0) | 76.3 (70.1-81.5) | 24.39 (19.4-30.1) | 54.4 (45.8-62.8) |
| **Residency** |  |  |  |  |  |  |  |  |
| Urban | 6761 | 1441 | 21.3 (19.9-22.8) | 52.2 (48.4-56.0) | 34.9 (31.2-38.8) | 66.8 (62.2-71.2) | 20.2 (16.8-24.0) | 57.8 (51.1-64.2) |
| Rural | 3304 | 578 | 17.5 (16.1-19.0) | 45.3 (41.4-49.1) | 23.8 (20.5-27.5) | 52.6 (47.1-58.0) | 12.6 (10.2-15.4) | 52.8 (45.0-60.4) |
| **Education** |  |  |  |  |  |  |  |  |
| No education | 3141 | 935 | 29.8 (27.8-31.8) | 50.6 (46.6-54.5) | 33.4 (29.6-37.3) | 65.9 (61.2-70.4) | 17.3 (14.5-20.6) | 52.0 (45.1-59.0) |
| Primary | 3122 | 572 | 18.3 (16.6-20.2) | 51.4 (46.4-56.3) | 28.8 (24.4-33.8) | 56.1 (48.9-63.2) | 16.8 (12.8-21.6) | 58.2 (48.1-67.7) |
| Secondary | 3291 | 438 | 13.3 (11.8-14.9) | 45.2 (39.8-50.8) | 29.6 (24.5-35.3) | 65.4 (56.7-73.2) | 18.3 (14.1-23.4) | 61.7 (51.6-71.0) |
| Higher | 511 | 74 | 14.4 (10.9-18.7) | 66.5 (52.8-77.8) | 45.9 (33.3-59.0) | 69.0 (53.1-81.5) | 33.8 (21.6-48.7) | 73.7 (53.0-87.5) |
| **Marital Status** |  |  |  |  |  |  |  |  |
| Married (Currently) | 7256 | 1568 | 21.6 (20.3-22.9) | 50.8 (47.6-54.0) | 31.9 (28.8-35.2) | 62.8 (58.5-67.1) | 17.9 (15.2-21.0) | 56.0 (50.0-61.9) |
| Never married (Single) | 1982 | 91 | 4.5 (3.6-5.8) | 21.7 (14.4-33.1) | 6.6 (2.7-15.3) | 30.5 (12.8-56.9) | 4.6 (1.4-13.5) | 69.4 (27.1-93.2) |
| Widowed/divorced | 827 | 360 | 43.6 (39.7-47.6) | 54.9 (48.4-61.2) | 31.9 (28.8-35.2) | 62.8 (58.5-67.1) | 21.8 (16.7-27.9) | 58.6 (48.2-68.3) |
| **Body Mass Index (BMI)** |  |  |  |  |  |  |  |  |
| <18.5 | 1437 | 190 | 13.2 (11.4-15.3) | 37.4 (29.9-45.7) | 18.6 (12.7-26.2) | 49.6 (36.6-62.3) | 7.9 (4.5-13.5) | 42.8 (27.2-60.0) |
| 18.5 – 24.9 | 5825 | 917 | 15.7 (14.6-16.9) | 42.5 (38.6-46.5) | 27.6 (23.6-31.2) | 64.0 (58.2-69.2) | 16.3 (13.0-20.2) | 60.0 (51.6-67.9) |
| 25 – 29.9 | 2142 | 654 | 30.5 (28.6-32.9) | 57.6 (52.8-62.3) | 37.3 (32.7-42.0) | 64.7 (59.0-70.1) | 20.2 (16.5-24.6) | 54.3 (46.4-61.9) |
| ≥30 | 661 | 258 | 39.0 (34.4-43.7) | 68.3 (60.7-75.1) | 43.3 (36.2-50.7) | 63.4 (54.3-71.5) | 25.6 (19.2-33.1) | 59.0 (47.5-69.6) |
| **Household wealth quintile (by residence)** |  |  |  |  |  |  |  |  |
| Poorest | 1862 | 350 | 18.7 (16.6-21.0) | 31.3 (26.3-36.8) | 14.8 (11.0-19.7) | 47.4 (38.2-56.8) | 9.3 (6.3-13.5) | 62.5 (47.8-75.2) |
| Poorer | 1929 | 340 | 17.6 (15.7-19.7) | 40.5 (34.7-46.5) | 23.8 (18.7-29.7) | 58.8 (49.3-67.6) | 12.8 (8.6-18.8) | 54.1 (40.0-67.8) |
| Middle | 2089 | 417 | 19.9 (17.7-22.3) | 53.6 (46.6-58.4) | 27.8 (22.5-33.9) | 52.9 (44.5-61.2) | 14.9 (10.9-20.0) | 53.6 (42.3-64.5) |
| Richer | 1916 | 371 | 19.4 (16.9-22.0) | 55.2 (48.7-61.5) | 33.6 (27.7-40.0) | 60.8 (52.7-68.3) | 17.4 (13.0-22.8) | 51.8 (41.1-62.4) |
| Richest | 2269 | 541 | 23.8 (21.2-26.7) | 63.3 (58.2-68.2) | 49.3-43.4-55.3) | 77.9 (71.1-83.4) | 29.6 (23.9-36.0) | 60.0 (52.1-67.4) |
| **Ecological zone** |  |  |  |  |  |  |  |  |
| Mountain | 552 | 111 | 20.0 (14.9-26.4) | 35.1 (22.2-50.6) | 12.5 (5.7-25.3) | 35.6 (22.5-51.3) | 6.2 (3.2-11.7) | 49.8 (27.2-72.5) |
| Hill | 4137 | 866 | 20.9 (19.3-22.6) | 52.0 (47.5-56.5) | 33.2 (29.1-37.6) | 63.9 (59.3-68.3) | 20.1 (16.4-34.4) | 60.4 (53.3-67.2) |
| Terai | 5375 | 1043 | 19.4 (17.9-21.0) | 50.3 (46.1-54.6) | 32.5 (28.2-37.1) | 64.5 (58.6-70.0) | 17.5 (13.9-21.8) | 53.8 (45.5-61.9) |
| **Province** |  |  |  |  |  |  |  |  |
| Bagmati | 2202 | 491 | 22.3 (19.5-25.4) | 61.4 (54.1-68.1) | 43.8 (36.8-51.0) | 71.3 (63.5-78.0) | 27.8 (20.8-36.1) | 63.5 (52.6-73.1) |
| Gandaki | 973 | 205 | 21.0 (18.7-23.5) | 51.2 (44.5-59.2) | 33.3 (25.6-42.1) | 64.2 (53.7-73.6) | 18.9 (12.6-27.4) | 56.7 (43.3-69.1) |
| Karnali | 556 | 79 | 14.2 (11.5-17.5) | 40.6 (31.4-50.5) | 15.8 (10.6-23.0) | 39.0 (27.6-51.8) | 10.1 (6.1-16.2) | 63.5 (43.4-79.7) |
| Koshi | 1840 | 470 | 25.5 (22.9-28.3) | 44.3 (39.1-49.8) | 24.6 (19.2-30.9) | 55.4 (46.2-64.3) | 10.3 (6.8-15.2) | 41.8 (29.1-55.7) |
| Lumbini | 1767 | 314 | 17.8 (15.3-20.5) | 51.2 (44.4-57.9) | 33.6 (26.9-41.0) | 65.7 (56.6-73.8) | 20.8 (15.2-27.7) | 61.8 (49.2-73.0) |
| Madhesh | 1895 | 313 | 16.5 (14.2-19.0) | 50.9 (43.1-58.7) | 34.4 (27.7-41.8) | 67.6 (57.7-76.1) | 19.0 (13.6-25.8) | 55.1 (42.7-66.9) |
| Sudurpashchim | 832 | 148 | 17.8 (15.2-20.8) | 31.3 (24.4-39.1) | 11.0 (6.9-17.0) | 35.1 (23.7-48.5) | 5.0 (2.77-8.8) | 45.4 (25.3-67.1) |
| **Note:** |  |  |  |  |  |  |  |  |

### Statistical analysis

We accessed the household member-recode file with individual-level data from the Demographic and Health Surveys (DHS) program. We report survey-weighted estimates adjusted for enumeration areas (EAs) and for disproportionate sampling weight and non-response. Continuum performance indicators were estimated for the national sample by all independent variables defined for this study and for provinces stratified by sex. We estimated the prevalence of hypertension in the overall population, the prevalence of awareness/diagnosis among those with hypertension, the prevalence of treatment both among those with hypertension and those who were aware of their condition, and the prevalence of control both among those on treatment and among all those with hypertension along with 95% confidence intervals (95% CI), has been estimated. Logistic regression was used to assess the factors associated with each outcome above.

We plotted the proportion of participants with hypertension who reached each step of the care cascade in each province against the province’s GDP per capita (in 2022/23 international dollars) for the year of survey data collection. The analysis provides insights into health system performance relative to a province’s wealth, given that financing hypertension care may be more feasible in wealthier provinces than poorer ones. To assess the cumulative losses at each step, we then estimated the overall care cascade, keeping the denominator constant (population with hypertension) throughout. We included all explanatory variables and determined a priori as potential confounders in a multivariate model. All analyses were done using the Svy command in Stata software version 17 *(StataCorp, College Station, TX, USA)*.

## Results

### Sample characteristics

In this study, 10,021 unweighted participants (female = 5787, male=4235) were surveyed. For the present analysis, we excluded 18 females and 13 males due to missing BMI data. Therefore, the analysis included 9,990 unweighted observations, representing 10,065 participants (4,321 males and 5,744 females). The participants had a mean (SD) age of 39⸱7 (17⸱9) years.

Nationally, over two-thirds of the population resided in urban areas. More than half of the respondents were younger than 40, and nearly 90% were aged between 15 and 64. Approximately 70% of the participants were married, and around one-third had received no formal education; notably, the proportion of women without formal education was more than double that of men. Over half of the participants were residents of the Terai ecological zone, and over one-fifth were from Bagmati province. The mean (SD) systolic blood pressure was 114⸱4 (113⸱7-115⸱1) mm Hg for women and 119⸱9 (119⸱2-120⸱7) mm Hg for men. The mean (SD) diastolic blood pressure was 75⸱1 (74⸱6-75⸱6) mm Hg for women and 77⸱0 (76⸱5-77⸱5) mm Hg for men. Detailed characteristics of the study population, disaggregated by gender and place of residence, are presented in **Table *1***.

## National-Level Care Continuum

### Prevalence and socio-demographic predictors of hypertension

The national prevalence of hypertension was 20⸱0% (95% CI, 19⸱0%-21⸱2%), higher in urban areas (21⸱3%, 95% CI, 19⸱9%-22⸱8%) than rural areas (17⸱5%, 95% CI, 16⸱1%-19⸱0%). Men had a higher prevalence (22⸱9%, 95% CI, 21⸱4%-24⸱4%) than women (17⸱9%, 95% CI, 16⸱7%-19⸱2%), and prevalence increased with age, particularly among individuals aged ≥65 years (47⸱4%, 95% CI, 43⸱9%-51⸱0%). Individuals with no formal education (29⸱8%, 95% CI, 27⸱8%-31⸱8%), widowed/divorced status (43⸱6%, 95% CI, 39⸱7%-47⸱6%), BMI ≥30 (39⸱0%, 95% CI, 34⸱4%-43⸱7%), and higher wealth quintiles (23⸱8%, 95% CI, 21⸱2%-26⸱7%) had higher prevalence rates.

In multivariate analysis, sex, age, education, marital status, BMI, and residence significantly influenced the odds of hypertension. Urban residents had higher odds (aOR=1⸱37, 95% CI, 1⸱15-1⸱63) than rural residents, while men exhibited higher odds than women (aOR=1⸱60, 95% CI, 1⸱40-1⸱83). The odds of hypertension increased with age, particularly among individuals aged ≥65 years (aOR=8⸱75, 95% CI, 6⸱87-11⸱15). Higher education was associated with lower odds of hypertension (aOR = 0⸱60, 95% CI, 0⸱43-0⸱85). Widowed/divorced (aOR= 3⸱15, 95% CI, 2⸱23-4⸱43) and currently married (aOR= 1⸱91, 95% CI, 1⸱46-2⸱48) individuals had higher odds than those never married. Additionally, compared to individuals with a normal BMI (18⸱5-24⸱9), underweight individuals had lower odds (aOR=0⸱74, 95% CI, 0⸱60-0⸱92), whereas overweight (aOR=2⸱34, 95% CI, 2⸱01-2⸱72) and obese individuals (aOR=3⸱78, 95% CI, 2⸱90-4⸱93) demonstrated higher odds of hypertension **Table *2***.

### Prevalence and socio-demographic predictors of patients’ awareness of their hypertension (diagnosed hypertension)

Among individuals with hypertension, 50⸱2% (95% CI, 47⸱3%-53⸱2%) reported being diagnosed (“aware”). Diagnosis rates were higher among urban residents, females, older adults (≥65 years), those with higher education levels, widowed/divorced or married individuals, those with a BMI ≥30, and individuals in higher wealth quintiles **Table *2***.

In multivariate analysis, age, marital status, BMI, and wealth quintile were significantly associated with hypertension awareness. Awareness was significantly higher among older individuals (≥65 years: aOR=3⸱92, 95% CI, 2⸱66-5⸱77) compared to the youngest age group (15-39 years). Widowed/divorced (aOR= 2⸱25, 95% CI, 1⸱10-4⸱59) and currently married (aOR= 2⸱30, 95% CI, 1⸱17-4⸱59) individuals had higher odds of awareness than those never married. Overweight (aOR= 1⸱67, 95% CI, 1⸱29-2⸱15) and obese individuals (aOR= 2⸱76, 95% CI, 1⸱90-4⸱02) had higher odds of awareness than those with normal or underweight BMI. Additionally, individuals in the highest wealth quintile had greater odds of awareness (aOR=2⸱56, 95% CI, 1⸱70-3⸱84) compared to those in the lowest wealth quintile **Table *3***.

**Table 3:**
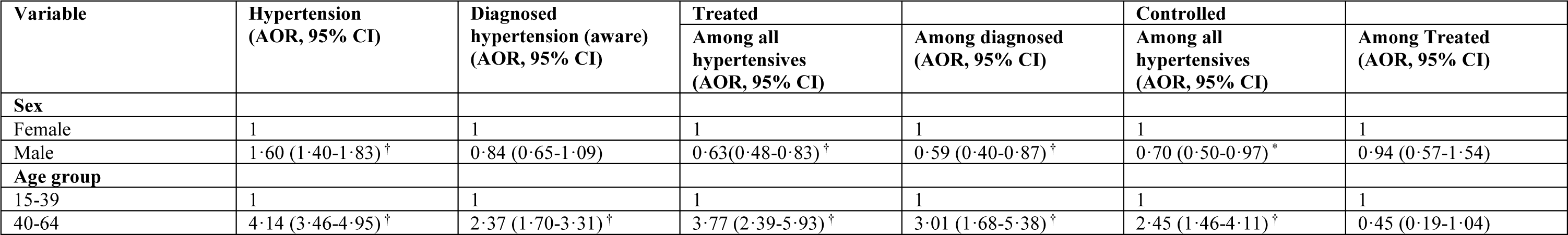
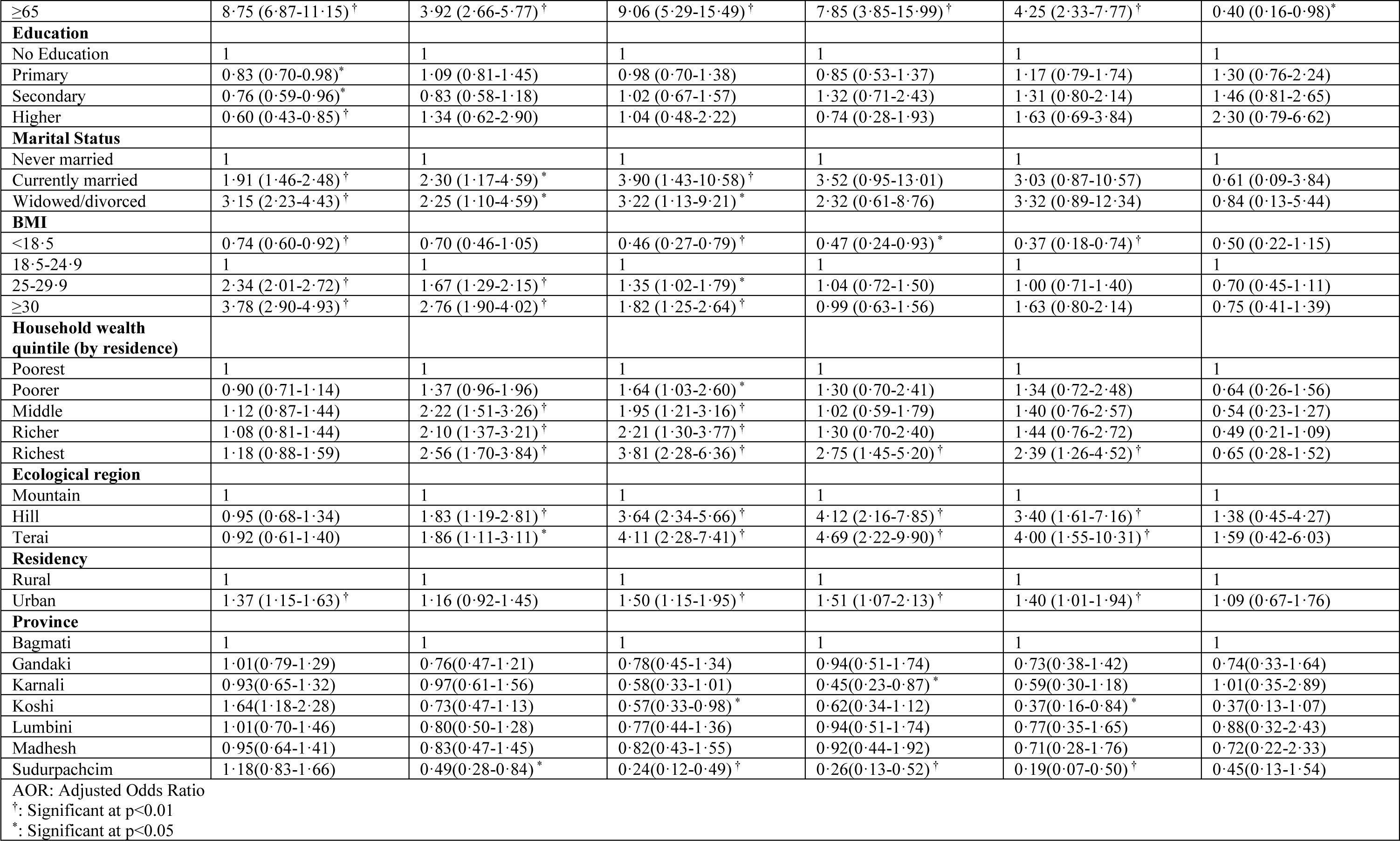
Adjusted analysis of factors associated with prevalence, diagnosis, treatment, and control of hypertension by socio-demographic factors in Nepal.

### Prevalence and socio-demographic predictors of treatment for hypertension

Among those diagnosed with hypertension, 63⸱2% (95% CI, 59⸱4%-66⸱8%) reported taking antihypertensive medication, representing 31⸱7% (95% CI, 28⸱8%-34⸱7%) of the overall hypertensive population. Treatment rates were higher among urban residents, females, older adults (≥65 years), individuals with higher education, widowed/divorced or married individuals, and those in higher wealth quintiles. These patterns were consistent among those diagnosed and treated for hypertension.

In multivariate analysis, among participants with hypertension, men were less likely to receive treatment (aOR=0⸱63, 95% CI, 0⸱48-0⸱83) than women⸱ Older individuals (≥65 years) were significantly more likely to receive treatment (aOR=9⸱06, 95% CI, 5⸱29-15⸱49) than the youngest age group (15-39 years). Compared to those in the lowest wealth quintile and rural residents, individuals in other health quintiles, particularly those in the highest wealth quintile (aOR= 3⸱81, 95% CI, 2⸱28-6⸱36) and urban residents (aOR= 1⸱50, 95% CI, 1⸱15-1⸱95) were more likely to receive treatment for hypertension. Widowed/divorced (aOR= 3⸱22, 95% CI, 1⸱13-9⸱21) and currently married (aOR= 3⸱90, 95% CI, 1⸱43-10⸱58) individuals had higher odds of receiving treatment among hypertensives **Table *3***.

### Prevalence and socio-demographic predictors of Controlled BP

Only 18⸱0% (95% CI, 15⸱5%-20⸱8%) of individuals with hypertension had controlled blood pressure, translating into 56⸱7% (95% CI, 51⸱2%-62⸱0%) among those receiving treatment. Among hypertensives, higher rates of controlled hypertension prevalence were observed in urban residents, females, older adults (≥65 years), individuals with higher education levels, widowed/divorced, those who were overweight/obese, and individuals in higher wealth quintiles. However, when considering only those receiving treatment the prevalence of controlled hypertension was higher among male, those with normal BMI, younger age group, participants with higher education.

In the multivariate analysis, men were less likely to achieve controlled blood pressure compared to women (aOR=0⸱70, 95% CI, 0⸱50-0⸱97). Among hypertensives, older participants (≥65; aOR=4⸱25, 95% CI, 2⸱33-7⸱77) were more likely to achieve blood pressure control than those aged 15-39. Additionally, participants with higher education levels (aOR=1⸱63, 95% CI, 0⸱69-3⸱84), those in the highest household wealth quintile (aOR=2⸱39, 95% CI, 1⸱26-4⸱52), and those living in urban areas (aOR=1⸱40, 95% CI, 1⸱01-1⸱94) had higher odds of achieving hypertension control compared to their counterparts. However, among those receiving treatment, the bivariate analysis ***(Supplementary Table 3)*** revealed that older participants (≥65) receiving treatment were less likely to achieve hypertension control (OR=0⸱46, 95% CI, 0⸱22-0⸱99), although this effect was not statistically significant in the multivariate analysis.

### Gaps in the hypertension care cascade

Among hypertensive patients, 50⸱2% (95% CI, 47⸱3%-53⸱2%) were aware of their diagnosis, 31⸱7% (95% CI, 28⸱8%-34⸱7%) were receiving treatment, and 18⸱0% (95% CI, 15⸱5%-20⸱8%) had controlled blood pressure. In men, 48⸱0% (95% CI, 44⸱1%-51⸱9%) were aware, 28⸱0% (95% CI, 24⸱6%-31⸱8%) were receiving treatment, and 16⸱4% (95% CI, 13⸱3%-20⸱1%) had controlled BP, corresponding to 58⸱6% (95% CI, 50⸱4%-66⸱3%) among those treated. Among women, 52⸱3% (95% CI, 48⸱8%-55⸱7%) were aware, 35⸱2% (95% CI, 31⸱5%-39⸱1%) were receiving treatment, and 19⸱5% (95% CI, 16⸱5%-22⸱9%) had controlled BP, corresponding to 55⸱3% (95% CI, 48⸱8%-61⸱5%) among those treated **Figure *1***.

### Province-Level Care Continuum

Among the seven provinces **Figure *2***, hypertension prevalence ranged from (14⸱2%, 95% CI, 11⸱5%-17⸱5%) in Karnali to (25⸱5%, 95% CI, 22⸱9%-28⸱3%) in Koshi. Notably, Bagmati had the highest rates of diagnosed hypertension (61⸱4%, 95% CI, 54⸱1%-68⸱1%), treated hypertension (43⸱8%, 95% CI, 36⸱8%-51⸱0%), and controlled hypertension (27⸱8%, 95% CI, 20⸱8%-36⸱1%). In contrast, Sudurpashchim had the lowest rates, with diagnosed hypertension at (31⸱3%, 95% CI, 24⸱4%-39⸱1%), treated hypertension at (11⸱0%, 95% CI, 6⸱9%-17⸱0%), and controlled hypertension at (5⸱0%, 95% CI, 2⸱7%-8⸱8%).

**Figure 2:**
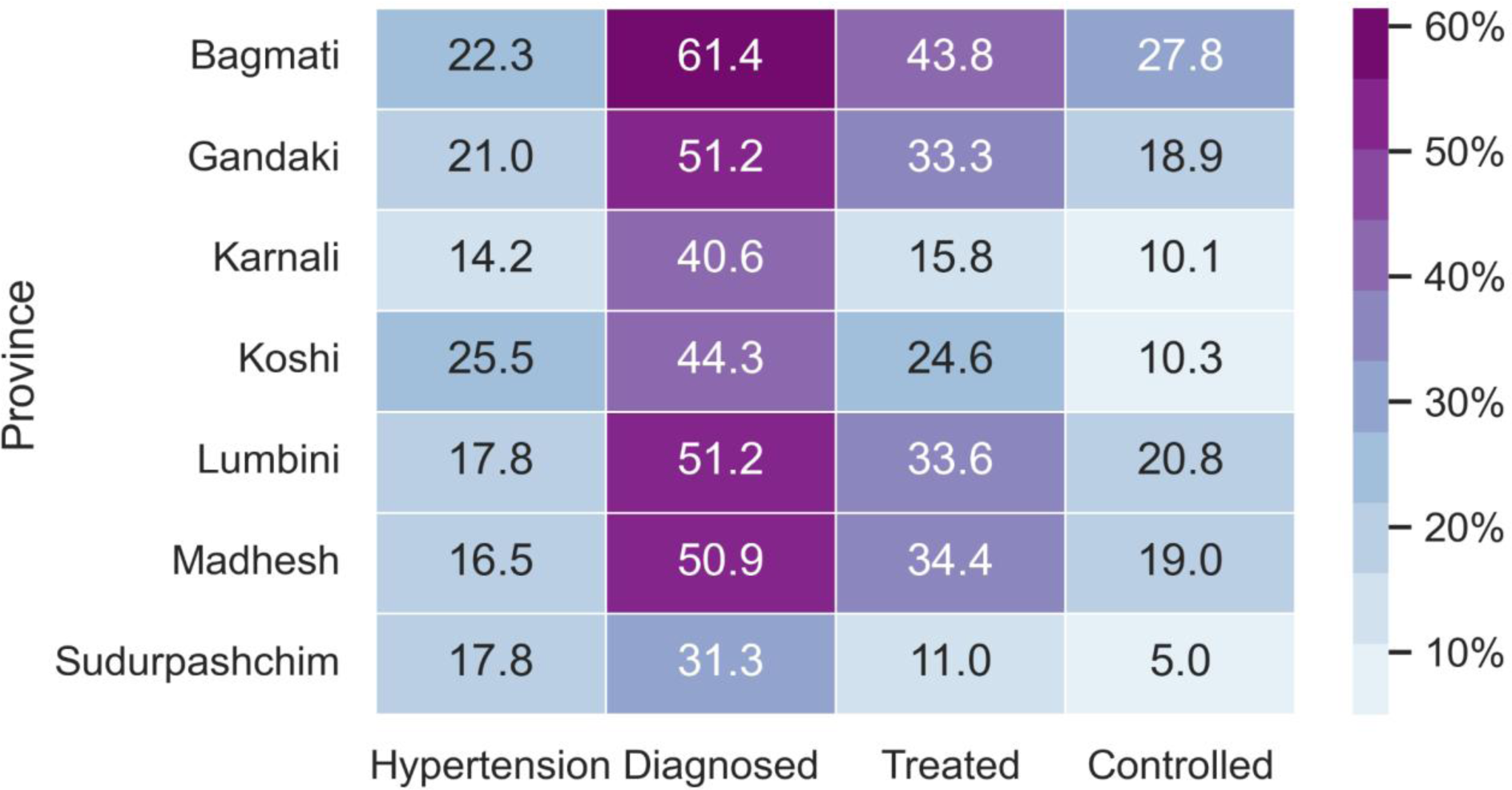
Heatmap for Care Continuum across provinces

In the multivariate analysis, after adjusting for all variables in **Table *2***, the prevalence of hypertension was significantly higher in Koshi province compared to Bagmati, which includes the capital city. Among all hypertensive participants, diagnosed hypertension was significantly less prevalent in Sudurpashchim, and treated and controlled hypertension rates were significantly lower in both Koshi and Sudurpashchim. Among diagnosed participants, treated hypertension was significantly lower in Karnali and Sudurpashchim. However, among treated participants, there was no significant difference in controlled hypertension prevalence across provinces.

Provinces with higher GDP per capita generally exhibited better performance on the hypertension cascade step ***(Figure 3 in Supplement)***. Notably, Lumbini and Madhesh provinces outperformed expectations in care cascade steps relative to their GDP per capita, while Sudurpashchim performed below predicted levels based on GDP per capita. The prevalence of hypertension was higher in urban areas compared to rural areas across all provinces ***(Figure 3 in Supplement 1)***. Urban areas also had higher rates of diagnosed and treated hypertension, except in Gandaki and Lumbini, and higher rates of controlled hypertension, except in Lumbini. Across all provinces, males had a higher prevalence of hypertension than females. Diagnosed hypertension was more prevalent in females, except in Sudurpashchim, Lumbini, and Gandaki. In Sudurpashchim and Karnali, females had lower rates of treated hypertension. Moreover, controlled hypertension was higher in females in most provinces, except Lumbini, Karnali, and Sudurpashchim.

## Discussion

This nationally representative study revealed that One-fifth of Nepalese adults aged 15 and above had hypertension, with an alarming 50% remaining undiagnosed. Of those diagnosed, only 63.2% reported taking medication, translating to merely 31.7% of all hypertensive individuals receiving treatment. Furthermore, a mere 18% of hypertensive adults overall and 56.7% of those treated had their blood pressure under control. Consequently, over 80% experienced gaps in the hypertension care continuum, being either undiagnosed, untreated, or treated but with uncontrolled hypertension, with the largest loss occurring at the diagnosis (50.2%) and treatment (18.5%) stages.

This study’s prevalence estimates for hypertension are lower than those reported for Nepal in 44 low- and middle-income countries (29.4%) (22) and the STEPS 2019 survey (25.4%) (23). However, they closely align with previous studies using NDHS 2016 data (24). These discrepancies in estimates may be attributed to variations in study populations and the restriction of the sample in these studies to adults aged 15–69 years. This study provides encouraging signs of progress in Nepal’s fight against hypertension. It reports the highest-ever national prevalence estimates for hypertension awareness, treatment, and control. Recent two nationally representative surveys conducted in 2016 (25) and 2019 (23)showed significantly lower rates, ranging from 20% to 40% for awareness, 10.3% to 20.2% for treatment, and a mere 3.8% to 10.5% for control. This positive trend might be due to the ongoing improvement in access to cardiovascular disease (CVD) services, as documented by the 2021 National Health Facility Survey (NHFS) (26). The NHFS revealed a surge in facilities offering CVD services, from 73% in 2015 to 90% in 2021, alongside an increase in the number of healthcare professionals who have recently completed CVD training and access to updated diagnosis and management guidelines. While further research is needed to confirm a causal relationship, these findings suggest a potential impact of strengthened healthcare facilities on tackling NCDs like hypertension in Nepal.

Substantial variability was observed in hypertension prevalence, diagnosis, treatment, and control across socio-demographic groups. Hypertension prevalence was higher among men in urban areas and marginally higher for women in rural areas. Women were more likely to be diagnosed and treated for hypertension, both among the hypertensive population and those already diagnosed. Furthermore, women had higher rates of controlled hypertension when considering all hypertensive patients. However, among those receiving treatment, the prevalence of controlled hypertension was lower in women, though this difference was not statistically significant. Overall, women performed better in the cascade of hypertension care than men. These findings are consistent with studies from 44 low- and middle-income countries (22), as well as other studies from Nepal (8,11,23) which indicate that being female is associated with higher rates of diagnosis, treatment, and control of hypertension. The NDHS 2016 from Nepal observed sex differences only in the treatment of hypertension (11), whereas STEPS 2019 observed sex differences in the diagnosis of hypertension but not in treatment or control after adjusting for other characteristics (23). The observed sex differences in the hypertension care continuum are likely due to higher healthcare-seeking behavior among women (27,28), as well as other social and biological factors that have been widely reported (29,30).

The proportions of individuals who received a diagnosis and were treated were lower among younger participants and those with lower household wealth. Therefore, understanding the unique healthcare perceptions and behaviors of young people (31) compared to older generations is crucial. Additionally, prioritizing interventions for those with lower wealth is essential, as this group faces significant barriers: limited access to high-quality care for cardiovascular events and a higher risk of financial hardship due to medical expenses. The country-level analysis revealed worse outcomes for each step of the care cascade (diagnosis, treatment, and control) among participants with lower BMI, those living in rural areas, and the Himalayan region. Low literacy levels and limited health services in rural Nepal likely contribute to lower awareness and treatment rates (32), while geographic difficulties exacerbate the disparity in access to healthcare in the Himalayan region. Individuals with higher BMI are likely to have more healthcare contact due to comorbidities (33,34), which explains their higher rates of hypertension screening and diagnosis. Surprisingly, the level of schooling did not significantly influence hypertension diagnosis, treatment, or control rates.

We found that 18% of hypertensive patients and 56.7% of those receiving treatment had controlled blood pressure. These rates were higher than those observed both nationally and in the subregion. A previous meta-analysis by Dhungana et al. (2016-2020) reported that only 38% (28–48%) of hypertensive patients and 8% (6–11%) of treated patients in Nepal had controlled blood pressure (8). Similarly, studies from India also revealed lower estimates for control rate (35,36). However, barriers to treatment success in the remaining patients should be investigated and addressed. One key barrier is the lack of national clinical guidelines for managing cardiovascular diseases, including hypertension potentially leading to inconsistent care and potentially suboptimal treatment plans. Another significant challenge is the unreliable supply of essential medications. Although some antihypertensives (amlodipine, enalapril, hydrochlorothiazide) are available at primary care centers, frequent stockouts (e.g., atenolol: about 70%, amlodipine: about 40% and thiazide diuretic: about 94% in 2021) pose a major hurdle (26). This inconsistency jeopardizes patient adherence and, ultimately, blood pressure control.

Among provinces, Bagmati Province, encompassing Kathmandu, Nepal’s capital city, has outperformed other provinces in the cascade of hypertension care. In contrast, Sudurpaschim Province, one of the least economically developed regions, performed the worst in managing hypertension care. This can be attributed to the significantly higher doctor-population density in the region. While Nepal faces a critical shortage of physicians, with a national health workforce index of 0.867 per 1,000 population, far below the WHO’s recommended 4.45 per 1,000 population, the doctor-population density in Kathmandu valley is estimated to be around 40 times higher than in rural areas of Nepal (37). Kathmandu valley’s concentrated healthcare resources likely improve the hypertension care cascade in Bagmati, with nearby rural areas also benefiting from the enhanced healthcare services available in the capital. However, Lumbini Province, despite having similar GDP, performs better than Sudurpashchim, suggesting that other factors likely contribute to the overall picture. Sudurpashchim’s struggles likely stem from its geographical remoteness and difficult terrain, hindering access to clinics (physical access). Additionally, lower insurance enrollment rates in the region (financial access) further restrict healthcare utilization (38). Existing data also reveals variations in health-seeking behaviors across provinces (39).

Overall, to improve the care continuum for hypertension in Nepal, our data are consistent with previous studies that suggest diagnosis is a critical step in realizing the downstream indicators, such as treatment and control. Several country-specific barriers to hypertension treatment and control have been identified at various levels: health system (lack of affordable services, resources), provider (inadequate counseling, long wait times, lack of guidelines, etc.), and patients (non-adherence, irregular follow-ups, lack of awareness, self-medication, lack of support, financial hardship etc.) (40). Addressing these multifaceted challenges in low- and middle-income countries (LMICs) like Nepal requires innovative strategies. The widespread use of mobile phones offers a unique opportunity. A pilot study in Nepal demonstrated that mobile interventions led to significant reductions in systolic and diastolic blood pressure compared to usual care [-7.09/-5.86 (p≤0.003) vs -0.77/-1.35 (p≥0.28) mmHg] (41), highlighting the potential of this approach to improve hypertension care in Nepal.

## Strength and limitations

This study has several strengths, including the use of the most recent nationally representative sample covering both urban and rural areas of all provinces in Nepal and appropriate statistical methods to estimate the weighted prevalence of hypertension. Additionally, the DHS employed validated tools and standard measurement techniques, such as appropriate cuff size selection, to minimize measurement errors.

However, the results should be considered in light of some limitations. The cross-sectional design with single-day blood pressure measurements restricts definitive diagnosis according to current guidelines, which recommend longitudinal monitoring (42). Additionally, our inclusion of potential confounders in multivariate analysis might not have captured all factors, and self-reported diagnosis and treatment introduce potential recall bias. Furthermore, our assessment focused solely on medication for hypertension control, neglecting lifestyle modifications. This likely underestimates the true prevalence of controlled hypertension, as participants who achieved control through lifestyle changes would not be classified as “controlled” in our study.

## Conclusion

In conclusion, about 1 in 5 Nepalese have hypertension, and over 80% of adults with hypertension are either undiagnosed, untreated, or treated but with uncontrolled hypertension. However, these national figures mask substantial provincial and socioeconomic disparities, with loss to hypertension care ranging from 70% in Bagmati to 95% in Sudurpaschim province and 91% among the poorest households compared to 70% in the wealthiest. While improvements are needed across the entire hypertension care cascade, the most significant gains lie in early diagnosis and ensuring access to affordable treatment, particularly for low-income households, remote areas, and younger populations who could benefit from low-cost medication programs.

## Authors’ contributions

Conceptualization: Ashok Khanal, Irusha Dahal, Sulochan GC Data curation: Ashok Khanal

Formal analysis: Ashok Khanal, Sulochan GC

Findings Interpretation: Ashok Khanal, Irusha Dahal, Sulochan GC, Sumi Mishra

Writing-original draft: Ashok Khanal, Irusha Dahal, Sulochan GC

Writing-review & editing: Rakesh Ghimire, Sharada Prasad Wasti, Vijay S. GC

All authors read and approved of the final manuscript.

## Ethics approval

This study was based on the analysis of deidentified, secondary data that are publicly available. We obtained permission to use the data from the ICF International/Demographic Health Survey (DHS) team.

## Data availability

Data used in this study are available from NDHS (2022) upon request.

## Data Availability

Data used in this study are available from NDHS (2022) upon request.

https://dhsprogram.com/data/dataset/Nepal_Standard-DHS_2022.cfm?flag=0

## Acknowledgments

This study was carried out using the datasets of Nepal Demographic Health Survey (NDHS) 2016. Hence, the authors of this study are thankful to DHS programme for offering the datasets

