## Supplementary Table 1 for "Hypertension care cascade in Nepal: findings from Nepal Demographic and Health Survey 2022"

**Supplementary appendix**

**Supplementary Table 1: Definitions of disease and care cascade of hypertension**

| **Term** | **Study Population (Denominator)** | **Ascertainment** | **Definitions** | **Categories** |
| --- | --- | --- | --- | --- |
| Hypertension | Analytic sample | The average of the last two measurements | An SBP ≥140 mm Hg or DBP ≥90 mm Hg or a person is taking any prescribed drugs to control raised blood pressure | Dichotomous (Yes, No) |
| Diagnosis | “Hypertension” as Yes | Self-reported | Answered “Yes” to: Ever been diagnosed by doctor/health worker with hypertension? | Dichotomous (Yes, No) |
| Treatment among hypertensive | “Hypertension” as Yes | Self-reported.  No information on dosage and type of medication used was obtained | “Answered “Yes” to: Are you taking medication to control blood pressure? | Dichotomous (Yes, No) |
| Treatment among diagnosed | “Diagnosis” as Yes | Self-reported.  No information on dosage and type of medication used was obtained | “Answered “Yes” to: Are you taking medication to control blood pressure? | Dichotomous (Yes, No) |
| Control among hypertensive | “Hypertension” as Yes | The average of the last two measurements | Blood pressure in no hypertensive range (<140/90mmHg) | Dichotomous (Yes, No) |
| Control among treated | “Treatment” as Yes | The average of the last two measurements | Blood pressure in no hypertensive range (<140/90mmHg) | Dichotomous (Yes, No) |

**Supplementary Table 2: Description of study variables**

| **Study variables** | **Ascertainment** | **Definitions** | **Categories** |
| --- | --- | --- | --- |
| Age | Self-reported | Age of the respondents | Ordinal  (15‐39; 40‐64; ≥65) |
| Sex | Self-reported | Gender of the respondents | Dichotomous  (Male, female) |
| Education | Self-reported | Education level of the respondents | Ordinal  [No education  Basic education (1-8^th^ grade)  Secondary education (9-12^th^ grade)  Higher education (Above 12^th^ grade)] |
| Body Mass Index (BMI) | Physical measurement | BMI of the respondents (kg/m2). Obtained by dividing weight (in kilograms) with square of the height (in meters) | Ordinal  [<18.5 (Underweight)  18.5 – 24.9 (Normal weight  25 – 29.9 (Overweight)  ≥30 (Obese)] |
| Marital Status | Self-reported | Marital status of the respondents | Polychotomous  [Single (Never Married)  Currently Married  Formerly Married (Divorced/Widowed)] |
| Household Wealth Quintile (urban- and rural-specific) | Principal component analysis | Composite index of household ownership of selected assets; obtained by principal component analysis | Polychotomous  [Q1 (Lowest)  Q2 (Second)  Q3 (Middle)  Q4 (Fourth)  Q5 (Highest)] |
| Place of residence | Place of interview | Whether the person is living in a rural or urban area at the time of interview | Dichotomous  (Urban, rural) |
| Ecological zone | Ecological zone of interview | Based on climate and landform Nepal is divided into three ecological zones | Polychotomous  (Mountain, Hill, Terai) |
| Province | Province of interview. | Province of residence. Province is the largest administrative unit in Nepal. | Polychotomous  (Bagmati, Gandaki, Karnali, Koshi, Lumbini, Madhesh, Sudurpashchim) |

**Supplementary Table 3: Unadjusted analysis of factors associated with prevalence, diagnosis, treatment, and control of hypertension by socio-demographic factors in Nepal**

| **Variable** | **Hypertension**  **(UOR, 95% CI)** | **Diagnosed**  **(UOR, 95% CI)** | **Treated** | | **Controlled** | |
| --- | --- | --- | --- | --- | --- | --- |
|  |  |  | **Among hypertensives**  **(UOR, 95% CI)** | **Among diagnosed**  **(UOR, 95% CI)** | **Among hypertensives**  **(UOR, 95% CI)** | **Among treated**  **(UOR, 95% CI)** |
| **Sex** |  |  |  |  |  |  |
| Female | 1 | 1 | 1 | 1 | 1 | 1 |
| Male | 1⸱35 (1⸱22-1⸱50) ^†^ | 0⸱84 (0⸱68-1⸱02) | 0⸱71 (0⸱58-0⸱88) ^†^ | 0⸱68 (0⸱51-0⸱90) ^†^ | 0⸱81 (0⸱62-1⸱06) | 1⸱14 (0⸱78-1⸱67) |
| **Age group, years** |  |  |  |  |  |  |
| 15-39 | 1 | 1 | 1 | 1 | 1 | 1 |
| 40-64 | 6⸱08 (5⸱26-7⸱03) ^†^ | 2·45 (1⸱80-3⸱33) ^†^ | 4⸱09 (2⸱77-6⸱04) ^†^ | 3⸱20 (1⸱89-5⸱39) ^†^ | 2⸱72 (1⸱69-4⸱39) ^†^ | 0⸱50 (0⸱23-1⸱09) |
| ≥64 | 11⸱55 (9⸱65-13⸱83) ^†^ | 3⸱08 (2⸱21-4⸱29) ^†^ | 6⸱67 (4⸱44-10⸱01) ^†^ | 6⸱14 (3⸱48-10⸱85) ^†^ | 3⸱81 (2⸱38-6⸱11) ^†^ | 0·46 (0·22-0·98) ^*^ |
| **Education** |  |  |  |  |  |  |
| No Education | 1 | 1 | 1 | 1 | 1 | 1 |
| Primary | 0⸱52 (0⸱46-0⸱60) ^†^ | 1⸱03 (0⸱81-1⸱30) | 0⸱80 (0⸱63-1⸱03) | 0⸱66 (0⸱46-0⸱92) | 0⸱96 (0⸱69-1⸱33) | 1⸱28 (0⸱82-2⸱01) |
| Secondary | 0⸱36 (0⸱30-0⸱42) ^†^ | 0⸱80 (0⸱61-1⸱05) | 0⸱83 (0⸱61-1⸱14) | 0⸱97 (0⸱64-1⸱48) | 1⸱06 (0⸱74-1⸱51) | 1⸱49 (0⸱93-2⸱38) |
| Higher | 0⸱39 (0⸱28-0⸱54) ^†^ | 1⸱95 (1⸱08-3⸱46) ^*^ | 1⸱69 (0⸱98-2⸱91) | 1⸱15 (0⸱56-2⸱33) | 2⸱43 (1⸱28-4⸱63) ^†^ | 2⸱59 (0⸱99-6⸱75) |
| **Marital Status** |  |  |  |  |  |  |
| Never married | 1 | 1 | 1 | 1 | 1 | 1 |
| Currently married | 5⸱75 (4⸱50-7⸱34) ^†^ | 3⸱73 (2⸱06-6⸱73) ^†^ | 6⸱61 (2⸱58-16⸱91) ^†^ | 3⸱84 (1⸱27-11⸱63) ^*^ | 4⸱52 (1⸱37-14⸱88) ^*^ | 0⸱56 (0⸱09-3⸱44) |
| Widowed/divorced | 16⸱16 (11⸱96-21⸱81) ^†^ | 4⸱39 (2⸱35-8⸱20) ^†^ | 8⸱33 (3⸱11 (-22⸱30) ^†^ | 4⸱77 (1⸱51-15⸱03) ^†^ | 5⸱78 (1⸱70-19⸱65) ^†^ | 0⸱62 (0⸱10-3⸱88) |
| **BMI** |  |  |  |  |  |  |
| <18⸱5 | 0⸱81 (0⸱64-0⸱99) ^†^ | 0⸱80 (0⸱55-1⸱18) | 0⸱60 (0⸱37-0⸱98) ^*^ | 0⸱55 (0⸱30-0⸱99) ^*^ | 0⸱44 (0⸱22-0⸱84) ^*^ | 0⸱49 (0⸱23-1⸱07) |
| 18⸱5-24⸱9 | 1 | 1 | 1 | 1 | 1 | 1 |
| 25-29⸱9 | 2⸱35 (2⸱06-2⸱68) ^†^ | 1⸱83 (1⸱46-2⸱31) ^†^ | 1⸱58 (1⸱25-2⸱01) ^†^ | 1⸱03 (0⸱74-1⸱42) | 1⸱29 (0⸱95-1⸱75) | 0⸱78 (0⸱52-1⸱19) |
| ≥30 | 3⸱42 (2⸱75-4⸱25) ^†^ | 2⸱91 (2⸱02-4⸱19) ^†^ | 2⸱04 (1⸱44-2⸱87) ^†^ | 0⸱97 (0⸱63-1⸱48) | 1⸱75 (1⸱12-2⸱73) ^*^ | 0⸱95 (0⸱54-1⸱68) |
| **Household wealth quintile (by residence)** |  |  |  |  |  |  |
| Poorest | 1 | 1 | 1 | 1 | 1 | 1 |
| Poorer | 0⸱92 (0⸱76-1⸱12) | 1⸱49 (1⸱05-2⸱10) ^*^ | 1⸱78 (1⸱14-2⸱79) ^*^ | 1⸱58 (0⸱92-2⸱68) | 1⸱44 (0⸱77-2⸱66) | 0⸱70 (0⸱30-1⸱65) |
| Middle | 1⸱07 (0⸱88-1⸱31) | 2⸱43 (1⸱73-3⸱41) ^†^ | 2⸱21 (1⸱42-3⸱41) ^†^ | 1⸱24 (0⸱76-2⸱04) | 1⸱71 (0⸱98-2⸱96) | 0⸱69 (0⸱32-1⸱48) |
| Richer | 1⸱04 (0⸱83-1⸱30) | 2⸱70 (1⸱87-3⸱90) ^†^ | 2⸱89 (1⸱84-4⸱55) ^†^ | 1⸱71 (1⸱02-2⸱86) ^*^ | 2⸱05 (-1⸱17-3⸱58) ^*^ | 0⸱64 (0⸱30-1⸱34) |
| Richest | 1⸱35 (1⸱10-1⸱67) ^†^ | 3⸱79 (2⸱74-5⸱24) ^†^ | 5⸱58 (3⸱69-8⸱44) ^†^ | 3⸱90 (2⸱33-6⸱54) ^†^ | 4⸱11 (2⸱47-6⸱83) ^†^ | 0⸱90 (0⸱45-1⸱77) |
| **Ecological region** |  |  |  |  |  |  |
| Mountain | 1 | 1 | 1 | 1 | 1 | 1 |
| Hill | 1⸱05 (0⸱72-1⸱53) | 2⸱00 (1⸱02-3⸱90) | 3⸱47 (1⸱42-8⸱49) ^†^ | 3⸱20 (1⸱61-6⸱35) ^†^ | 3⸱77 (1⸱81-7⸱86) ^†^ | 1⸱53 (0⸱56-4⸱21) |
| Terai | 0⸱95 (0⸱65-1⸱39) | 1⸱87 (0⸱96-3⸱63) | 3⸱36 (1⸱38-8⸱15) ^†^ | 3⸱28 (1⸱65-6⸱53) ^†^ | 3⸱18 (1⸱52-6⸱66) ^†^ | 1⸱17 (0⸱42-3⸱27) |
| **Residency** |  |  |  |  |  |  |
| Rural | 1 | 1 | 1 | 1 | 1 | 1 |
| Urban | 1⸱27 (1⸱11-1⸱45) ^†^ | 1⸱32 (1⸱06-1⸱64) ^*^ | 1⸱71 (1⸱32-2⸱21) ^†^ | 1⸱81 (1⸱34-2⸱44) ^†^ | 1⸱75 (1⸱27-2⸱42) ^†^ | 1⸱122 (0⸱81-1⸱84) |
| UOR: Unadjusted Odds Ratio  ^†^: Significant at p<0.01  ^*^: Significant at p<0.05 |  |  |  |  |  |  |

**Supplementary Table 4: Hypertension prevalence stratified by gender and place of residence across Provinces in Nepal**

| **Province** | **Total population** | **Number of individuals with hypertension** | **Hypertension**  **(%, 95% CI)** | **Diagnosed**  **(%, 95% CI)** | **Treated** | | **Controlled** | |
| --- | --- | --- | --- | --- | --- | --- | --- | --- |
|  |  |  |  |  | **Among Hypertension**  **(%, 95% CI)** | **Among Diagnosed**  **(%, 95% CI)** | **Among Hypertension**  **(%, 95% CI)** | **Among Treated**  **(%, 95% CI)** |
| Koshi Urban | 1186 | 309 | 26⸱0 (22⸱6-29⸱7) | 45⸱2 (38⸱1-52⸱5) | 27⸱9 (20⸱6-36⸱6) | 61⸱7 (49⸱4-72⸱6) | 11⸱6 (6⸱9-18⸱8) | 41⸱7 (25⸱8-59⸱6) |
| Koshi Rural | 654 | 161 | 24⸱6 (20⸱7-28⸱9) | 42⸱6 (35⸱3-50⸱4) | 18⸱2 (11⸱8-27⸱2) | 42⸱8 (30-56⸱6) | 7⸱7 (3⸱9-14⸱6) | 42⸱2 (25⸱2-61⸱3) |
| Madhesh Urban | 1382 | 247 | 17⸱9 (14⸱8-21⸱4) | 52⸱65 (42⸱8-62⸱3) | 36⸱1 (27⸱9-45⸱2) | 68⸱6 (56⸱7-78⸱4) | 20⸱5 (13⸱9-29⸱1) | 56⸱8 (42⸱0-70⸱4) |
| Madhesh Rural | 513 | 65 | 12⸱7 (10⸱5-15⸱4) | 44⸱4 (35⸱6-53⸱7) | 28⸱0 (20⸱8-36⸱6) | 63⸱1 (46⸱7-77⸱0) | 13⸱2 (7⸱7-21⸱1) | 47⸱0-28⸱5-66⸱2) |
| Bagmati Urban | 1744 | 395 | 22⸱6 (19⸱2-26⸱4) | 65⸱2 (56⸱6-72⸱9) | 49⸱4 (50⸱0-58⸱0) | 75⸱8 (66⸱4-83⸱2) | 31⸱5 (22⸱9-41⸱5) | 63⸱6 (51⸱8-74⸱0) |
| Bagmati Rural | 456 | 96 | 21⸱0 (17-26) | 45⸱6 (35⸱8-55⸱7) | 20⸱4 (14⸱2-28⸱5) | 44⸱9 (34⸱9-55⸱3) | 12⸱6 (7⸱5-20⸱4) | 61⸱9 (37⸱0-81⸱8) |
| Gandaki Urban | 667 | 143 | 21⸱4 (18⸱4-24⸱7) | 49⸱8 (40⸱1-59⸱5) | 33⸱1 (22⸱9-45⸱3) | 66⸱5 (51⸱4-78⸱9) | 19⸱7 (11⸱2-32⸱3) | 59⸱6 (41⸱1-75⸱6) |
| Gandaki Rural | 306 | 61 | 20⸱1 (16⸱7-24⸱0) | 56⸱7 (46⸱7-66⸱2) | 33⸱8 (24⸱8-44⸱1) | 59⸱6 (46⸱0-71⸱9) | 16⸱9 (11⸱4-24⸱4) | 50⸱1 (34⸱4-65⸱9) |
| Karnali Urban | 303 | 51 | 16⸱7 (12⸱6-21⸱8) | 48⸱4 (35⸱4-61⸱6) | 16⸱9 (10⸱3-25⸱5) | 35⸱0 (22⸱9-49⸱5) | 12⸱6 (6⸱9-21⸱9) | 74⸱2 (44⸱0-91⸱3) |
| Karnali Rural | 253 | 29 | 11⸱2 (8⸱1-15⸱4) | 26⸱7 (17⸱2-38⸱9) | 13⸱9 (6⸱4-27⸱5) | 52⸱0 (26⸱5-76⸱5) | 5⸱6 (2⸱1-13⸱7) | 40⸱2 (16⸱5-69⸱5) |
| Lumbini Urban | 976 | 195 | 20⸱0 (16⸱2-24⸱4) | 50⸱0 (40⸱9-59⸱0) | 32⸱4 (23⸱2-43⸱0) | 64⸱8 (51⸱3-76⸱2) | 20⸱43 (12⸱9-30⸱8) | 63⸱1 (44⸱7-78⸱2) |
| Lumbini Rural | 792 | 119 | 15⸱1 (12⸱2-18⸱2) | 53⸱1 (42⸱9-63⸱1) | 35⸱7 (26⸱6-45⸱9) | 67⸱2 (54⸱9-77⸱5) | 21⸱4 (14⸱4-30⸱5) | 59⸱9 (42⸱5-75⸱1) |
| Sudurpashchim Urban | 503 | 101 | 20⸱1 (17⸱0-23⸱6) | 31⸱44 (23⸱2-41⸱0 | 12⸱9 (7⸱3-21⸱9) | 41⸱2 (25⸱8-58⸱7) | 5⸱2 (2⸱4-11⸱1) | 40⸱4 (18⸱4-67⸱1) |
| Sudurpashchim Rural | 329 | 47 | 14⸱3 (10⸱0-20⸱2) | 31⸱1 (19⸱1-46⸱2) | 6⸱8 (3⸱2-13⸱8) | 22⸱0 (10⸱4-40⸱6) | 4⸱5 (2⸱0-9⸱8) | 65⸱7 (18⸱5-94⸱2) |
| Koshi Female | 1038 | 257 | 24⸱7 (21⸱9-27⸱9) | 46⸱2 (39⸱1-53⸱5) | 28⸱1 (21⸱1-36⸱3) | 60⸱8 (49⸱2-71⸱3) | 12⸱3 (8⸱3-17⸱7) | 43⸱7 (30⸱3-58⸱1) |
| Koshi Male | 802 | 213 | 26⸱5 (23⸱0-30⸱2) | 42⸱1 (33⸱9-50⸱7) | 20⸱3 (13⸱4-29⸱5) | 48⸱3 (35⸱1-61⸱7) | 7⸱8 (3⸱5-16⸱3) | 38⸱7 (19⸱2-62⸱6) |
| Bagmati Female | 1199 | 245 | 20⸱4 (16⸱8-24⸱5) | 66⸱3 (56⸱9-74⸱6) | 48⸱9 (39⸱4-58⸱5) | 73⸱7 (63⸱4-82⸱0) | 28⸱8 (20⸱6-38⸱6) | 58⸱9 (47⸱5-69⸱4) |
| Bagmati Male | 1003 | 246 | 24⸱5 (21⸱2-28⸱2) | 56⸱5 (47⸱5-65⸱0) | 38⸱7 (30⸱1-48⸱0) | 68⸱5 (56⸱6-78⸱3) | 26⸱8 (18⸱1-37⸱6) | 69⸱3 (52⸱2-82⸱3) |
| Madhesh Female | 1115 | 134 | 12⸱0 (9⸱9-14⸱6) | 58⸱8 (49⸱0-68⸱0) | 43⸱5 (34⸱6-52⸱8) | 74 (62⸱9-82⸱6) | 26⸱0 (17⸱2-37⸱3) | 59⸱8 (41⸱4-75⸱8) |
| Madhesh Male | 780 | 178 | 22⸱8 (19⸱2-26⸱9) | 45⸱0 (35⸱4-54⸱9) | 27⸱5 (20⸱1-36⸱5) | 61⸱3 (47⸱5-73⸱4) | 13⸱6 (8⸱4-21⸱2) | 49⸱5 (34⸱5-64⸱5) |
| Gandaki Female | 562 | 108 | 19⸱2 (16⸱8-21⸱9) | 49⸱7 (37⸱5-62⸱0) | 36⸱0 (25⸱8-47⸱7) | 72⸱4 (60⸱4-81⸱8) | 20⸱6 (13⸱1-31⸱0) | 57⸱3 (40⸱7-72⸱3) |
| Gandaki Male | 411 | 96 | 23⸱4 (19⸱5-27⸱9) | 54⸱2 (43⸱5-64⸱7) | 30⸱3 (21⸱9-40⸱3) | 55⸱9 (40⸱9-69⸱9) | 16⸱9 (10⸱3-26⸱6) | 55⸱9 (37⸱9-72⸱5) |
| Lumbini Female | 1003 | 177 | 17⸱6 (15⸱3-20⸱1) | 50⸱7 (42⸱0-59⸱4) | 35⸱3 (27⸱4-44⸱0) | 69⸱6 (59⸱0-78⸱4) | 20⸱5 (14⸱0-28⸱8) | 58 (41⸱3-73⸱0) |
| Lumbini Male | 764 | 138 | 18⸱0 (14⸱4-22⸱3) | 51⸱7 (42⸱8-60⸱6) | 31⸱5 (23⸱9-40⸱1) | 60⸱9 (48⸱6-71⸱9) | 21⸱2 (14⸱9-29⸱2) | 67⸱3 (50⸱1-80⸱8) |
| Karnali Female | 334 | 40 | 11⸱9 (9⸱1-15⸱4) | 42⸱3 (29⸱8-55⸱8) | 12⸱4 (7⸱5-19⸱8) | 21⸱3 (18⸱1-43⸱7) | 6⸱0 (2⸱6-13⸱5) | 48⸱7 (18⸱7-79⸱6) |
| Karnali Male | 222 | 39 | 17⸱7 (13⸱7-22⸱5) | 38⸱9 (28⸱7-50⸱2) | 19⸱4 (10⸱2-33⸱5) | 49⸱7 (27⸱6-71⸱9) | 14⸱1 (6⸱7-27⸱4) | 73⸱1 (47⸱2-89⸱2) |
| Sudurpashchim Female | 493 | 70 | 14⸱2 (11⸱7-17⸱2) | 27⸱7 (18⸱4-39⸱3) | 9⸱4 (4⸱9-17⸱2) | 34⸱0 (18⸱1-54⸱6) | 4⸱2 (1⸱7-9⸱8) | 44⸱4 (18⸱8-73⸱4) |
| Sudurpashchim Male | 339 | 78 | 23⸱0 (18⸱8-27⸱9) | 34⸱6 (26⸱5-43⸱7) | 12⸱4 (7⸱2-20⸱5) | 35⸱9 (21⸱5-53⸱5) | 5⸱74 (2⸱8-11⸱4) | 46⸱1 (20⸱2-74⸱2) |

**Supplement Figure 1: Process of selection of participants and analytical sample**

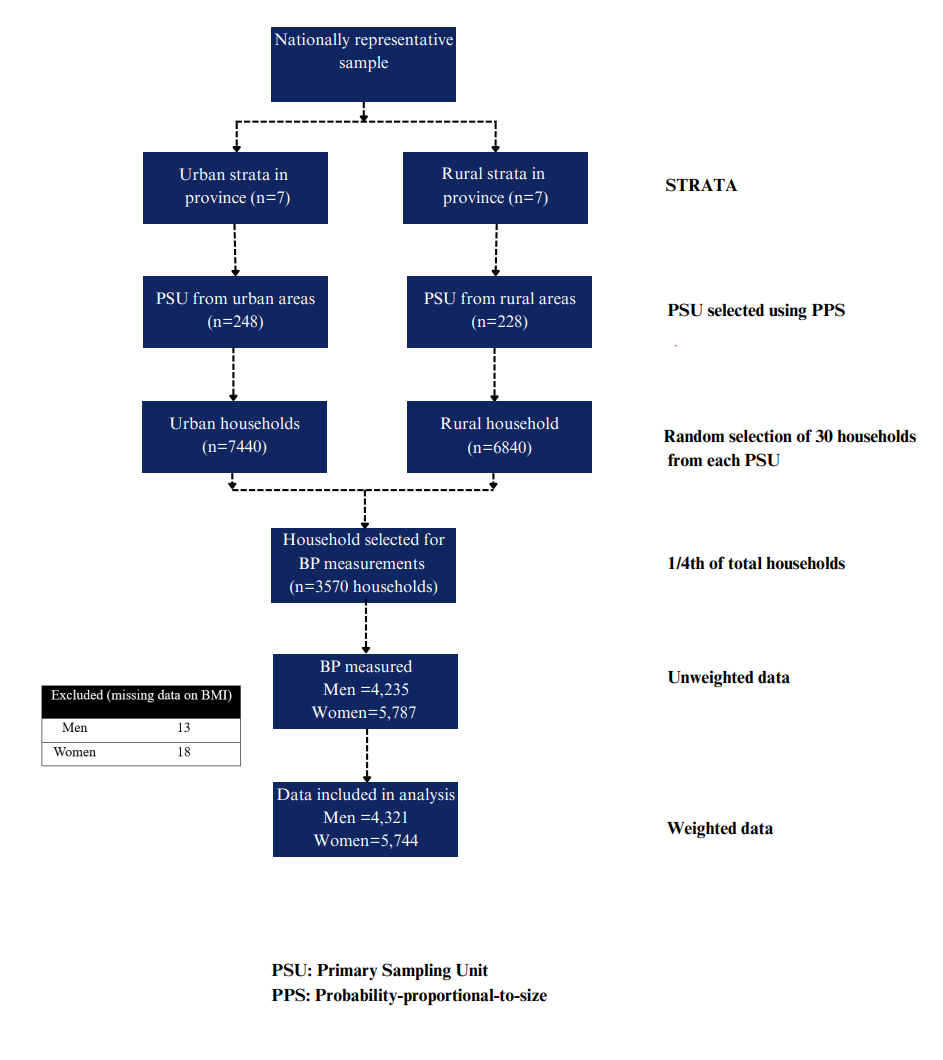

**Supplement Figure 2: Heatmap for Care Continuum across provinces by residency and sex**

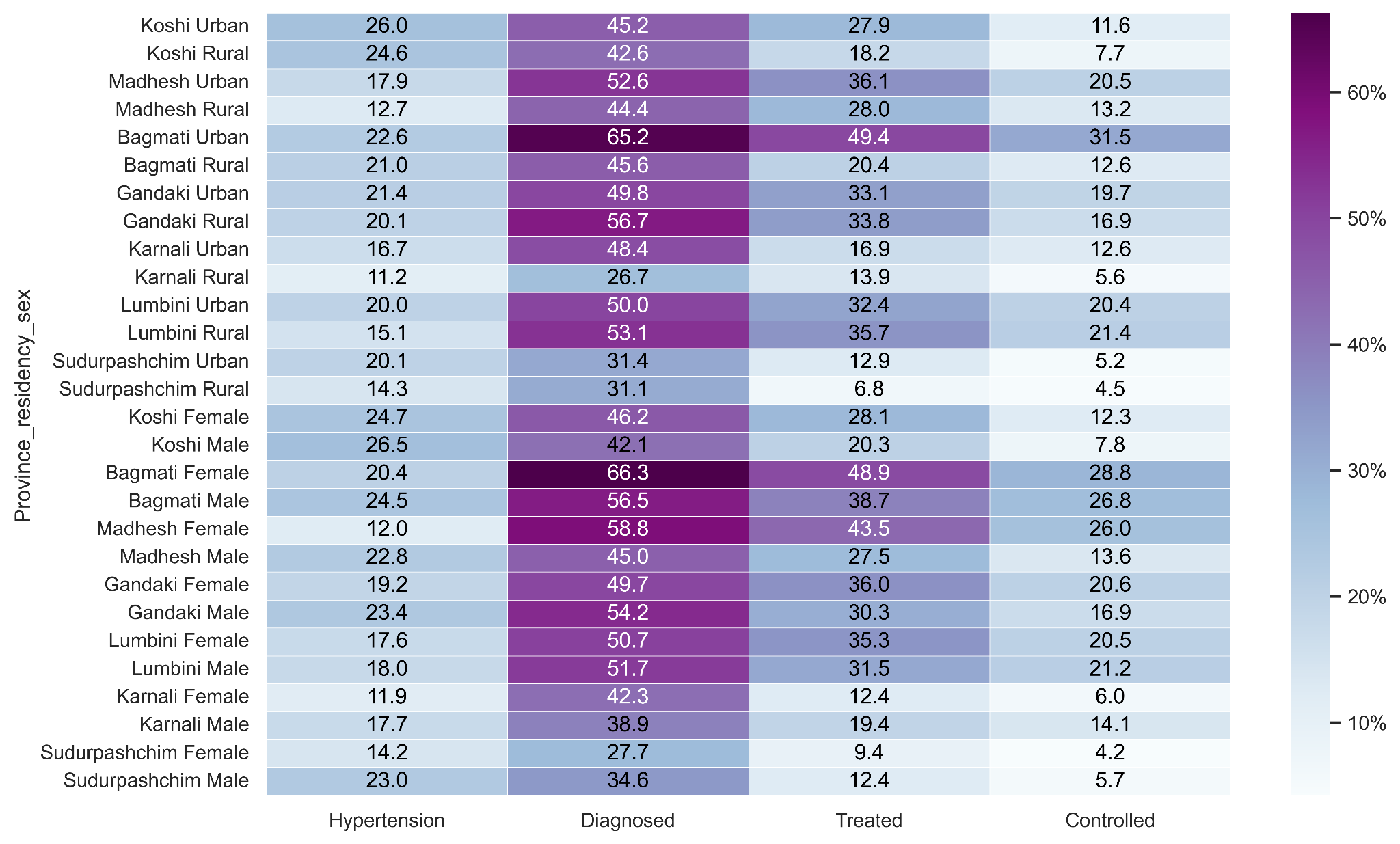

**Supplement Figure 3: The association between the province's gross domestic product (GDP) and each care cascade step.**

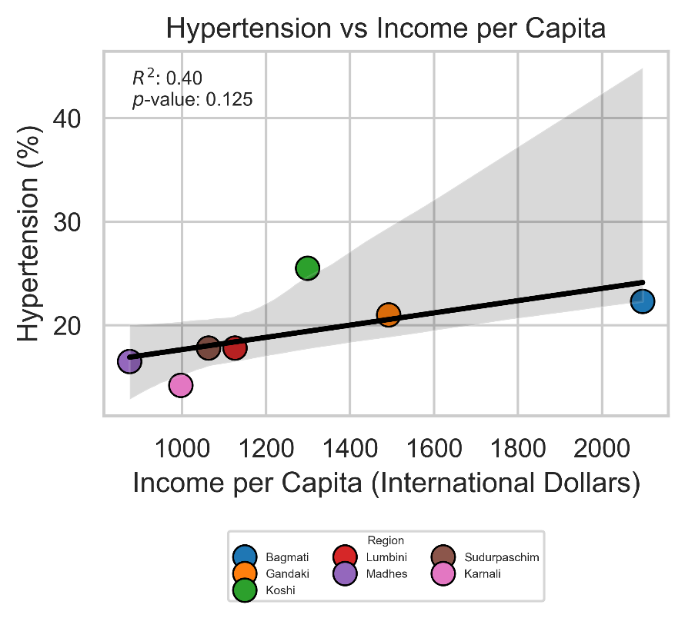

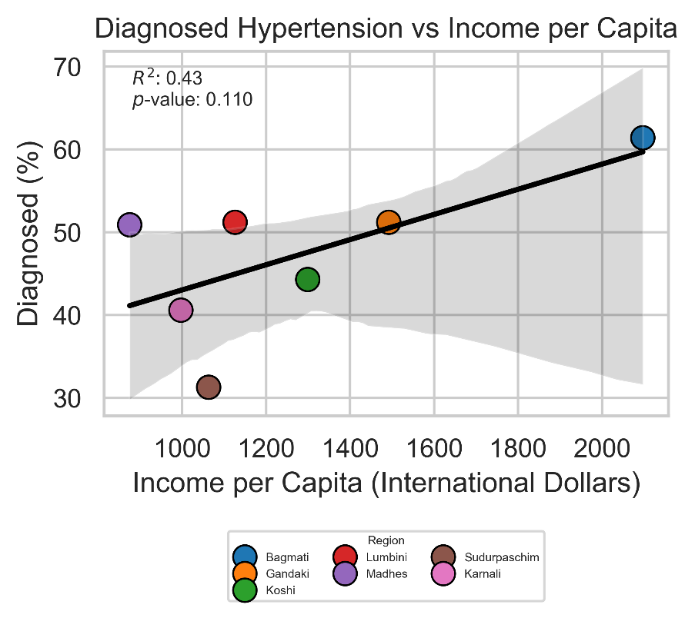

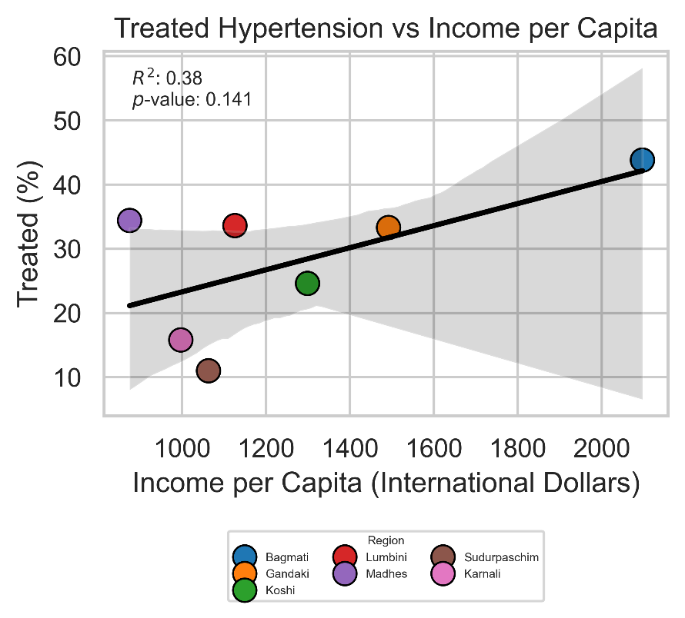

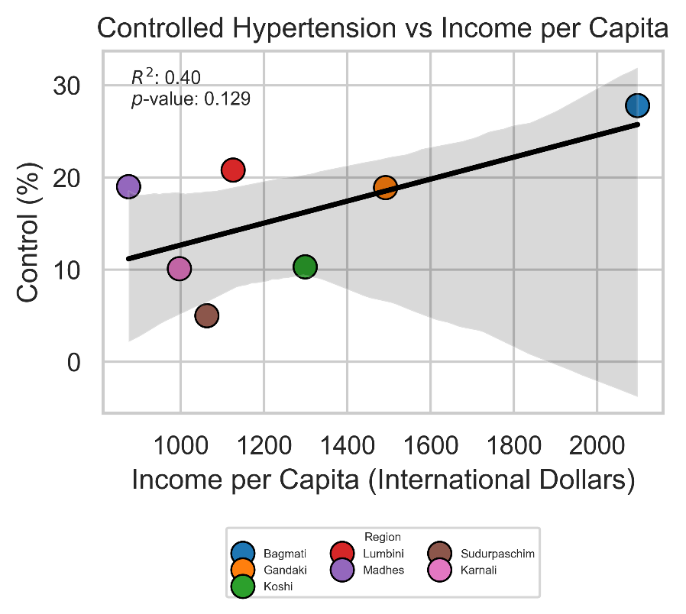

Province GDP data (for 2022/23) were obtained from the National Statistics Office (NSO), office of the Prime Minister and Council of Ministers of. The grey ribbon depicts the pointwise 95% prediction interval. The vertical bars depict 95% confidence intervals. The figure shows p values, which refer to the regression coefficient of the ordinary least squares regression line (with each state having the same weight). Similarly, R2 values are for the ordinary least squares regression of the province' cascade achievement onto their GDP per capita.
